# Pre-conceptional maternal vitamin B12 supplementation improves offspring neurodevelopment at 2 years of age: PRIYA trial

**DOI:** 10.1101/2021.09.09.21263316

**Authors:** Naomi D’souza, Rishikesh V Behere, Bindu Patni, Madhavi Deshpande, Dattatray Bhat, Aboli Bhalerao, Swapnali Sonawane, Rohan Shah, Rasika Ladkat, Pallavi Yajnik, Kalyanaraman Kumaran, Caroline Fall, Chittaranjan S Yajnik

## Abstract

**Background:** Nutritional interventions during the first 1000 days of life improves lifelong health. Better pre-conceptional maternal nutrition improves the nutrition of the early embryo. Vitamins B12 and folate are important for fetal neural development. Vitamin B12 deficiency is common in India.

**Methods:** In the Pune Rural Intervention in Young Adolescents (PRIYA) adolescents (N=557, 226 females) were provided with vitamin B12 (2µg/day) with or without multiple micronutrients, or a placebo, from preconception until delivery. All groups received mandatory iron and folic acid. We used the Bayley’s Scale of Infant Development (BSID-III) at 24-42 months of age to investigate effects on offspring neurodevelopment. We examined cord blood concentrations of brain-derived neurotropic factor (BDNF).

**Results:** Participants in the three groups had similar baseline B12 levels. These improved in the B12 supplemented groups at pre-conceptional and pregnancy (28 weeks gestation) measurements, reflected in higher cord holo-TC levels compared to the placebo. Neurodevelopmental outcomes are available for 74 children. Offspring in the B12 alone group (n=21) performed better than the placebo (n=27) on cognition (p=0.044) and language (p=0.020) domains (adjusted for maternal baseline B12 levels). There were no differences between the B12+MMN (n=26) and placebo group. Cord blood BDNF levels were highest in the B12 alone group (not statistically significant).

**Conclusion:** Pre-conceptional vitamin B12 supplementation improved maternal B12 status and offspring neurodevelopment at 2 years of age. The usefulness of cord BDNF as a marker of brain development needs further investigation. Our results highlight the importance of intervening in the pre-conceptional period.

## 1.0 Introduction

The developing fetus is dependent on its mother for its nutrition. Maternal undernutrition before and during pregnancy affects fetal growth and development, the effects of which may predispose an individual to undesirable outcomes in later life. This concept is called ‘fetal programming’. This is the backbone for the Developmental Origins of Health and Disease (DOHaD) paradigm (1,2). Pregnancy and the first two years of life (1000 days) are considered the most crucial window for programming (3).

Maternal nutritional factors (both macro and micronutrients) influence neurodevelopmental processes *in utero*, such as neurogenesis, myelination, synaptogenesis, and cortical brain growth (3).Vitamins B12 and folate are of special interest due to their effects on cellular growth (DNA synthesis, and epigenetic regulation by methylation) mediated by the one carbon metabolism pathway (4). Offspring of mothers with low maternal vitamin B12 and folate during pregnancy have a higher risk of neural tube defects and neurodevelopmental disorders (autism, ADHD), poorer cognitive development and smaller brain volumes in childhood (5–8). In animal models (rats), offspring of mothers exposed to a high folate and low vitamin B12 diet show lower levels of the neurotrophic factor Brain Derived Neurotrophic Factor (BDNF) in the brain, and poorer cognitive function (9,10).

In India, vitamin B12 deficiency is widely prevalent in pregnant women (50-70%) (11,12) and is attributable to the socio-cultural practice of vegetarianism and poor economic status (13–16). This deficiency is associated with a range of adverse pregnancy and offspring health outcomes (17). In prospective birth cohorts (the Pune Maternal Nutrition study and the IAEA B12 study) from western India, we have earlier shown that exposure to low maternal vitamin B12 *in utero* is associated with poorer cognitive functioning at the age of 2 and 9 years in the offspring (18,19). However, public health policy in India mandates only iron and folic acid supplementation to women in the reproductive age group, and during pregnancy and lactation. A randomized controlled trial in South India showed that supplementing 50 µg/day oral B12 from 14 weeks of pregnancy until 6 weeks postpartum improved B12 concentrations in breast milk, the vitamin B12 status of infants at 6 weeks and infant cognitive function at 30 months of age (20,21).

Important milestones in fetal neural development such as neural tube closure are completed by 26-28 days of gestation (22). The majority of pregnancies in India are unplanned, and by the time pregnancy is detected (typically between 10-14 weeks gestation) this early developmental window is lost. Pre-conceptional intervention will ensure that the mother has improved vitamin stores during the early neurodevelopmental period. The success of pre-conceptional folic acid supplementation in preventing neural tube defects is well known (23–25). Few studies have examined the effects of pre-conceptional maternal micronutrient supplementation on offspring neurodevelopment in India. This approach will expand the 1000 days concept to include the pre-conceptional period.

The Pune Rural Intervention in Young Adolescents (PRIYA) is a pre-conceptional vitamin B12 and multi micronutrient supplementation trial in adolescent participants of the Pune Maternal Nutrition Study. We report neurodevelopmental outcomes at 2 years of age in the offspring of female participants of the trial. We hypothesised that pre-conceptional B12 supplementation in the mothers would contribute to better neurodevelopmental outcomes in their offspring.

## 2.0 Material and Methods

### 2.1 PRIYA trial

The PRIYA trial methods have been previously published (26). Briefly, The Pune Maternal Nutrition Study (PMNS) is a pre-conceptional observational birth cohort set up in 1993 (Figure 1). Non-pregnant women were recruited from six villages around Pune and those who became pregnant were followed through pregnancy. Seven hundred and sixty-two children were born and followed up serially. At age ∼17 years, 690 participants from the PMNS cohort (11) were screened for inclusion in the PRIYA trial. Of these, 133 were excluded due to severe vitamin B12 deficiency or systemic illnesses. Five hundred and fifty-seven (266 females) participants were randomized (Figure 1) to receive either a placebo, B12 (2µg/day) + multiple micronutrients (MMN) or B12 alone (2µg/day). The composition of the MMN tablet was guided by the WHO/UNICEF/UNU international multiple micronutrient preparation (UNIMMAP), excluding folic acid because the mandated IFA tablets (100 mg elemental Iron and 500 µg folic acid once a week) were given to all participants as per Government of India recommendations. Female participants in the trial received supplementation daily until their first delivery. They and the study team were blinded to the vitamin/micronutrient supplementation.

**Figure 1.**
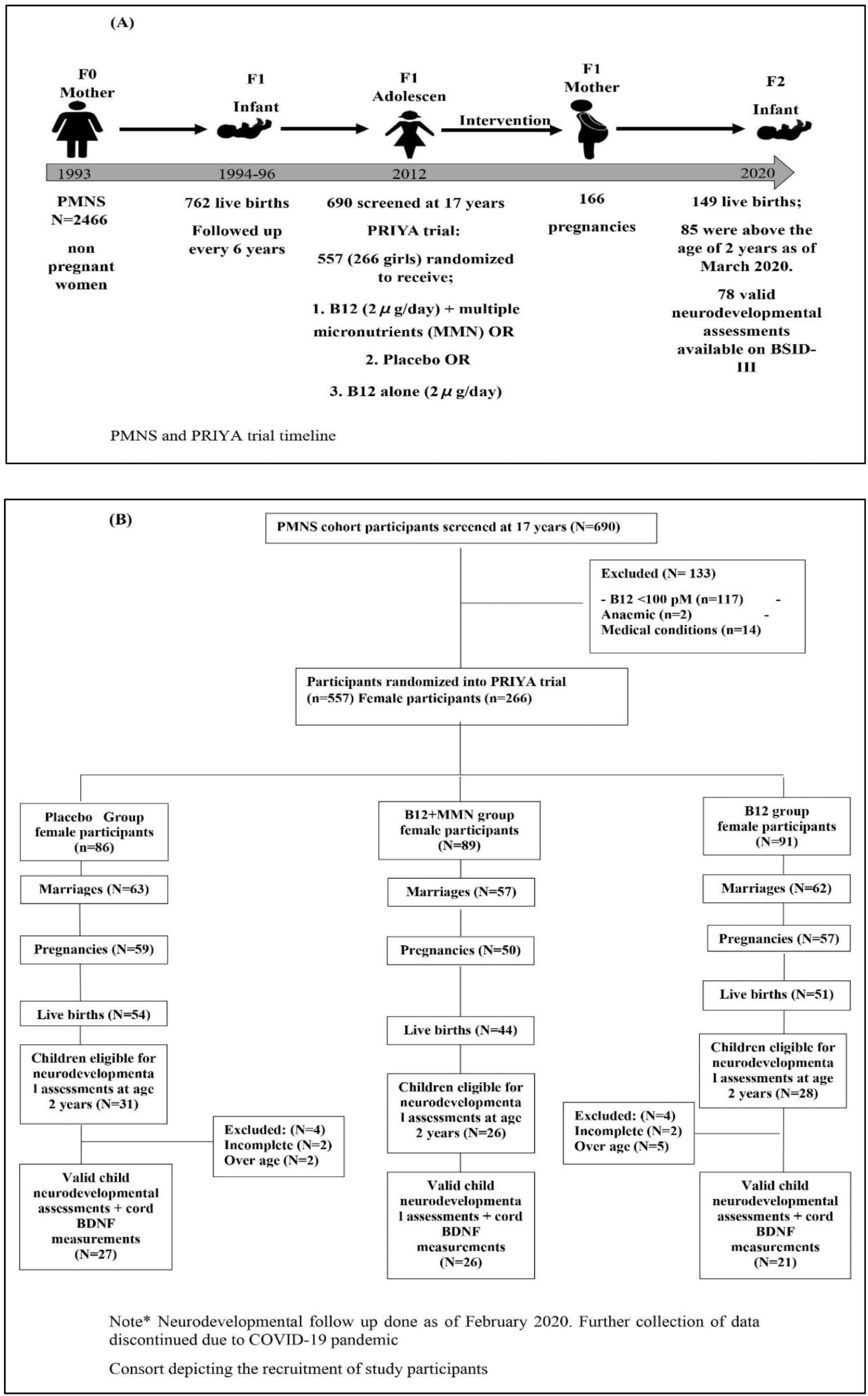
Study timeline and consort

Participants were followed up regularly for any health problems, and marriages were recorded. Married women were monitored to detect pregnancy which was confirmed by a urine pregnancy test. At 28 weeks gestation, mothers visited the Diabetes Unit, KEM Hospital Research Center, Pune for evaluation. This included socio demographic information (assessed using the Standard of Living Index questionnaire of the NFHS of India), anthropometry, physical examination and blood measures of circulating vitamins B12, folate, B2 and B6. At birth, delivery details and the baby’s size were recorded. Maternal and cord blood samples were collected.

These measurements were conducted in participants at baseline, 6-12 months after the start of the intervention (at ∼18 years of age), at 28 weeks gestation, and in cord blood. A Hemogram was measured on a Beckman Coulter analyzer (Miami, Florida, USA) on the day of the collection. Plasma vitamin B12 and folate were measured using a microbiological assay and total homocysteine, vitamin B2 and B6 by HPLC as described in the PRIYA trial methods publication (26). Plasma BDNF was measured in cord blood using ELISA (XpressBio) (Frederick, USA)

### 2.2 Neurodevelopmental assessments

The offspring born in the trial were followed up every 6 months until 2 years of age, for measurements of their growth. Once they reached 24 months of age, the parents were approached regarding participation in the neuro-cognitive study, and their written informed consent was obtained. An appointment was scheduled for the neurodevelopmental assessment at the Child Development Center (TDH center), KEM Hospital, Pune. The child and mother were transported to the center from their home on the morning of the assessment.

The neurodevelopmental assessment was performed using the Bayley’s Scale of Infant Development (BSID-III) (27). The BSID-III assesses the developmental status of infants from 1 to 42 months of age. The scales assess five domains across three main subscales: 1) cognitive - which includes items such as attention to familiar and unfamiliar objects, looking for a fallen object, and pretend play, 2) language - which assesses receptive and expressive language, via recognition of objects and people, following directions, and naming objects and pictures, and 3) motor - which assesses gross and fine motor skills such as grasping, sitting, stacking blocks, and climbing. The assessment was performed by trained clinical psychologists certified to perform the BSID-III. Testing was carried out in a quiet room, with a parent or guardian present, and instructions were provided in a language that was comfortable for the child. All children were assessed within a time window of 24 to 42 months of age. Each test protocol was independently reviewed and scored by two raters.

The BSID-III test yields raw scores based on the performance of the child on test items for cognitive, expressive, and receptive communication, and fine and gross motor skills. The raw scores were converted into age standardized scaled scores as recommended in the manual. Summation of the scaled scores yields 3 composite scores for the cognitive, language and motor skills domains. We used the composite scores in our analysis. Composite scores were categorized into average, below or above average performance, based on standardized criteria provided in the manual, where average is 100 with SD of 15 and a score of <85 is considered to be below average (27).

As part of ongoing assessments in the PMNS cohort, maternal IQ was assessed in some of the mothers at age 22∼24 years using the Weschler’s Adult Intelligence Scale-IV (WAIS-IV).

### 2.3 Ethical considerations

Details of community participation in the planning of this trial has been described earlier (26). The trial was approved by the KEM Hospital Research Centre Ethics committee (no 1242) and monitored by a Data Safety Monitoring Board (DSMB) and a Scientific Advisory Committee (SAC). The trial was registered with the CTRI (2012/12/003212) and ISRCTN (32921044). Neurodevelopmental assessment of the offspring was also approved by the Institutional ethics committee of the KEM Hospital research center and registered in (clinicaltrials.gov ID: NCT03088189).

### 2.4 Statistical Analysis

The purpose of our analysis was to see if pre-conceptional B12 and micronutrient supplementation in the mothers led to improvement in offspring neurodevelopmental performance (composite BSID-III scores) at 2 years of age. We also investigated the effect of intervention on circulating vitamin levels in the mother and cord blood, and on cord blood BDNF levels.

We first examined whether randomization had equally distributed potential confounders such as parental education and standard of living index, and maternal age, IQ, and anthropometry, across the three intervention groups.

All data were represented as either mean and standard deviation (for normally distributed variables) or median inter-quartile range (IQR, for skewed variables). All right skewed variables were log transformed. We used t-tests or ANOVA to test the significance of differences in outcomes between the intervention groups, and ANCOVA to test the significance of differences in outcomes between the intervention groups after adjusting for maternal B12 levels at screening. We used the Mann-Whitney U test to the significance of difference in cord BDNF values between the intervention groups.

## 3.0 Results

Of the 266 women randomized in the trial, 182 were married, 166 became pregnant, and 149 delivered a live baby. Between May 2017 and February 2020, we approached the parents of 85 children who had attained the age of 2 years of age, for participation in the neurodevelopmental study. Further assessments after February 2020 had to be halted due to the COVID-19 pandemic. None of the children had significant neurodevelopmental disorders (cerebral palsy, seizure disorders, or neural tube defects). Seven children who were above the inclusion age of 42 months as per the BSID norms, were excluded from analysis after confirming that they had achieved appropriate neurodevelopment for 42 months of age. Assessment could not be completed in 4 children. Our analysis is based on the remaining 74 children. The median age of the children at the time of performing the BSID was 29 months (Table1). There were 42 boys and 32 girls; of these, 27 were in the placebo group, 26 in the B12+MMN and 21 in the B12 alone group. There were no differences in gestational age at delivery, birth weight, length or head circumference amongst the offspring in the three intervention groups. Similarly, there were no differences in parental education, standard of living index, maternal age, or IQ. (Table 1)

**Table 1.** Maternal characteristics at baseline and pregnancy, and child characteristics

| Variables | n | Placebo Group | n | B12+MMN Group | n | B12 Group | p values |  |
| --- | --- | --- | --- | --- | --- | --- | --- | --- |
| Parental sociodemographic characteristics |  |  |  |  |  |  |  |  |
| Maternal age at 28 weeks gestation (years) | 25 | 19.8 (1.0) | 25 | 19.4 (1.1) | 21 | 19.7 (1.1) | 0.555 |  |
| Maternal education (years) | 26 | 12.5 (11.0,13.0) | 26 | 12.0 (10.0, 13.0) | 21 | 12.0 (11.0, 13.5) | 0.639 |  |
| Maternal height (cms) | 25 | 158.2 (5.2) | 25 | 158.7 (5.0) | 21 | 157.8 (4.8) | 0.852 |  |
| Maternal weight at 28 weeks gestation (kgs) | 25 | 55.4 (48.6, 59.3) | 25 | 51.2 (49.4, 54.4) | 21 | 52.9 (47.0, 60.7) | 0.656 |  |
| Maternal IQ | 21 | 76.6 (9.5) | 15 | 74.4 (8.8) | 17 | 75.8 (7.2) | 0.751 |  |
| Standard of Living Index | 26 | 36.0 (30.5, 40.5) | 26 | 38.0 (31.0, 40.0) | 21 | 37.0 (32.0, 40.0) | 0.923 |  |
| Paternal Education (years) | 25 | 14.0 (10.5, 15.0) | 24 | 12.0 (10.0, 15.0) | 19 | 12.0 (10.0, 15.0) | 0.656 |  |
| Maternal Micronutrients |  |  |  |  |  |  | p values (B12+MMN vs Placebo) | p values (B12 vs Placebo) |
| At screening |  |  |  |  |  |  |  |  |
| Vitamin B12 (pM) | 27 | 151.0 (122.0, 193.0) | 26 | 159.5 (134.0, 219.0) | 21 | 138.0 (125.0, 190.0) | 0.350 | 0.860 |
| Folate (nM) | 27 | 20.9 (15.3, 24.6) | 26 | 15.7 (11.3, 26.6) | 21 | 20.8 (15.3, 29.1) | 0.357 | 0.698 |
| Homocysteine (μmol/L) At 18 years | 27 | 20.1 (15.1, 38.0) | 26 | 18.6 (15.3, 30.3) | 21 | 27.5 (17.0, 39.6) | 0.646 | 0.434 |
| Vitamin B12 (pM) | 25 | 162.0 (125.9, 192.5) | 26 | 285.0 (205.8, 368.7) | 18 | 274.7 (224.7, 388.2) | <0.001*** | <0.001*** |
| Folate (nM) | 26 | 23.0 (17.2, 29.8) | 26 | 21.2 (15.3, 28.8) | 18 | 20.4 (14.9, 28.3) | 0.734 | 0.925 |
| Homocysteine (μmol/L) | 27 | 16.7 (11.7, 28.3) | 26 | 9.60 (8.30, 13.4) | 18 | 10.6 (9.22, 16.0) | <0.001*** | 0.013* |
| At 28 weeks gestation |  |  |  |  |  |  |  |  |
| Hemoglobin (gm/dl) | 25 | 10.4 (9.5, 11.0) | 25 | 10.2 (9.4, 11.0) | 21 | 10.4 (9.1, 10.7) | 0.491 | 0.638 |
| Vitamin B12 (pM) | 25 | 134.0 (95.5, 163.0) | 25 | 164.0 (149.0, 218.5) | 21 | 204.0 (173.5, 261.0) | 0.007** | <0.001*** |
| Holo-TC (pM) | 25 | 14.8 (8.85, 25.1) | 25 | 21.9 (15.3, 36.5) | 21 | 21.3 (16.9, 36.8) | 0.027* | 0.012* |
| Folate (nM) | 25 | 47.9 (18.0, 71.5) | 25 | 20.6 (10.2, 49.7) | 21 | 28.5 (16.6, 51.4) | 0.043* | 0.302 |
| Vitamin B2 (pM) | 25 | 244.0 (210.5, 273.0) | 25 | 276.0 (229.5, 304.5) | 20 | 244.0 (221.7, 269.5) | 0.028* | 0.852 |
| Viamin B6-pyridoxal-5-phospate (pM) | 24 | 3.5 (2.3, 4.6) | 25 | 4.6 (3.3, 7.4) | 21 | 3.1 (2.6, 4.8) | 0.117 | 0.357 |
| Vitamin B6-pyridoxal (pM) | 15 | 1.0 (0.8, 1.6) | 12 | 1.1 (0.9, 1.3) | 12 | 1.3 (1.0, 1.7) | 0.786 | 0.922 |
| Homocysteine (umol/L) | 25 | 7.0 (5.0, 9.2) | 25 | 6.3 (4.3, 8.1) | 21 | 5.1 (3.9, 7.2) | 0.559 | 0.550 |

| Child Characteristics |  |  |  |  |  |  |  |  |
| --- | --- | --- | --- | --- | --- | --- | --- | --- |
| Child age at assessment (months) | 27 | 27 (26, 34) | 26 | 29 (27, 36.2) | 21 | 29 (26, 32) | 0.623 | 0.901 |
| Gender | 27 | Boys=18 (66.7%) | 26 | Boys=13 (50%) | 21 | Boys=11 (52.3%) |  |  |
| Birth Anthropometry |  |  |  |  |  |  |  |  |
| Gestation age (weeks) | 27 | 39.0 (38.0, 40.2) | 26 | 39.0 (38.0, 40.2) | 21 | 39.4 (38.8, 40.2) | 0.920 | 0.936 |
| Birth weight (gm) | 27 | 2908.6 (412.5) | 26 | 2809.2 (458.6) | 21 | 2788.9 (315.9) | 0.411 | 0.277 |
| Birth length (cm) | 27 | 49.1 (46.8, 49.8) | 26 | 48.2 (47.4, 49.8) | 20 | 48.5 (47.2, 49.3) | 0.990 | 0.328 |
| Head circumference (cm) | 27 | 33.4 (1.0) | 26 | 33.1 (1.0) | 20 | 33.0 (0.9) | 0.237 | 0.142 |
| Cord Micronutrients |  |  |  |  |  |  |  |  |
| Vitamin B12 (pM) | 27 | 226.0 (138.0, 289.0) | 26 | 275.5 (181.7, 313.7) | 21 | 289.0 (167.0, 446.0) | 0.240 | 0.200 |
| Holo-TC (pM) | 27 | 40.7 (23.3, 81.9) | 26 | 79.4 (39.2, 125.0) | 21 | 96.1 (39.4, 125.0) | 0.021* | 0.048* |
| Folate (nM) | 27 | 55.9 (37.9, 70.8) | 26 | 52.0 (36.8, 68.1) | 21 | 42.7 (31.3, 80.0) | 0.473 | 0.278 |
| Vitamin B2 (pM) | 26 | 357 (73.7) | 25 | 316 (73.5) | 20 | 314 (67.3) | 0.053 | 0.924 |
| Vitamin B6-pyridoxal-5-phosphate (pM) | 12 | 29.3 (18.6, 42.7) | 13 | 25.0 (19.1, 39.5) | 13 | 17.5 (11.6, 43.9) | 0.494 | 0.298 |
| Vitamin B6-pyridoxal (pM) | 26 | 4.80 (3.6, 7.9) | 25 | 5.70 (4.6, 8.3) | 21 | 4.80 (3.2, 6.9) | 0.187 | 0.806 |
| Homocysteine (μmol/L) | 27 | 8.30 (6.8, 11.6) | 26 | 6.30 (4.8, 9.8) | 21 | 6.60 (4.6, 11.9) | 0.134 | 0.342 |
| BDNF (pg/ml) | 27 | 70.0 (31.0, 299.0) | 26 | 106.0 (31.0, 412.2) | 21 | 195.0 (31.0, 512.0) | 0.620 | 0.364 |

Values represented as Mean (SD), Median (25<sup>th</sup>, 75<sup>th</sup>) or n (%)
\*p<0.05, \*\*p<0.01 \*\*\*p<0.001 p-values calculated by t-test
Holo-TC, holotranscobalamin; BDNF, Brain Derived Neurotrophic Factor

The children who were not invited for the study because they were below 24 months of age differed from those studied; they had higher socio-economic status and parental education, higher maternal and cord B12 and holotranscobalamin (holo-TC) and lower cord homocysteine compared to the study group. (Supplementary Table 1).

### 3.1 Effect of supplementation on maternal and newborn micronutrient status, and birth measures

At baseline, maternal B12 and holo-TC levels were similar across the three intervention groups (Table 1). Fifty one percent of the females had vitamin B12 deficiency at screening (B12<150 pM), and this reduced to 22% at 6-12 months after starting intervention. There was a rise in vitamin B12 and holo-TC levels in the B12 supplemented groups compared to the placebo group, both pre-conceptionally (18 years of age) and at 28 weeks of gestation. Cord blood levels of holo-TC were significantly higher in the B12 supplemented groups compared to the placebo group, though B12 levels were similar.

Plasma homocysteine concentrations which were high but similar at baseline in the three groups of mothers, fell substantially in the vitamin B12 supplemented groups pre-conceptionally. During pregnancy, as expected, plasma homocysteine concentrations fell in all groups and were similar in the three groups during pregnancy as well as in cord blood.

Circulating folate concentrations were similar at baseline in the three groups and increased during pregnancy (due to supplementation). Folate levels were significantly lower at 28 weeks gestation in the B12+MMN group as compared to the placebo. Folate levels were similar in the cord blood across the groups. Circulating vitamin B2 and B6 concentrations increased from baseline in the B12 +MMN group before and during pregnancy and in cord blood.

Hemoglobin concentrations were similar in the mother and the baby across all the groups.

### 3.2 Comparison of BSID scores and cord BDNF between supplementation groups

Age standardized composite scores in the domains of cognition, motor and language development were obtained on 74 children. There was no difference in performance between males and females (supplementary Table 3). No significant developmental delays were observed in any of the children (score < 69). Few children showed a below average performance on the cognitive (4.0%, n=3), motor (4.0%, n=3), and language domain (8.1%, n=6) (score < 85) (supplementary table 2).

The offspring of mothers in the B12 alone group performed the best in the cognitive and language domains, and significantly better than the placebo group (Table 2). This difference persisted after adjusting for the baseline plasma vitamin B12 concentrations, the latter were not related to neurodevelopmental outcomes. There was a 5 -7% increase in cognition and language composite scores in the B12 alone group over the placebo group.

**Table 2.** Comparison between placebo and supplemented groups on BSID-III domains

| BSID-III domains | Placebo Group | B12+MMN Group | B12 Group | #p value | | \$p value | |
| --- | --- | --- | --- | --- | --- | --- | --- |
|  |  |  |  | Group (B12+MMN vs Placebo) | Group (B12 vs Placebo) | Group (B12+MMN vs Placebo) | Group (B12 vs Placebo) |
| **Cognitive** | 90.0<br>(85.0, 95.0) | 90.0<br>(85.0, 96.2) | 95.0<br>(90.0, 100) | 0.969 | 0.034* | 0.781 | 0.044* |
| **Motor** | 94.0<br>(91.0, 100.0) | 95.5<br>(90.2, 100.0) | 97.0<br>(91.0, 107.0) | 0.687 | 0.818 | 0.522 | 0.384 |
| **Language** | 92.2 (7.8) | 93.7 (9.87) | 98.6 (10.1) | 0.556 | 0.020* | 0.633 | 0.020* |

Values represented as Mean (SD) or Median (25<sup>th</sup>, 75<sup>th</sup>)
\*p<0.05

There were no significant differences between the B12+MMN group and the placebo group on any of the neurodevelopmental subscales.

The two intervention groups had higher cord BDNF values than the placebo group, the B12 alone group had the highest values, however the difference was non-significant (Table 1). Cord blood BDNF values did not show significant associations with any of the BSID-III composite scores.

## 4.0 Discussion

In this rural Indian population with a substantial prevalence of B12 deficiency, we found that supplementation of adolescents with 2 µg /day of B12 significantly improved their own B12 status (total B12 and holo-TC) and cord blood holo-TC. Offspring whose mothers received vitamin B12 alone performed better than offspring of mothers in the placebo group in neurodevelopmental assessments (cognitive and language domain of the BSID-III test at 24-42 months of age). Offspring whose mothers received B12+MMN performed similarly to the placebo group.

The role of pre-conceptional folic acid supplementation in preventing NTDs is well established, especially in western (mainly non-vegetarian) populations (23,24). In vegetarian populations like India, vitamin B12 is likely to play a similar role, probably because both folate and B12 act as cofactors for the enzyme methionine synthase, in methylation reactions. Studies in India have highlighted an association of both maternal vitamin B12 and folate with different outcomes in the offspring including neurodevelopmental performance. Studies in Pune showed an association of low maternal vitamin B12 status (low holo-TC concentrations and TCN2 polymorphisms) with risk of NTD, and a positive association between maternal vitamin B12 status during pregnancy and offspring neurocognitive performance at 2 and 9 years of age (5,18,19). A study in North Indian children aged 12–18 months found that both vitamin B12 and folate status had significant associations with cognitive performance (28) while a study in Mysore found that higher maternal folate concentrations, but not vitamin B12, during pregnancy were associated with better cognitive ability in children at 9-10 years of age (29). Adequate status of both vitamins is likely to be important for brain development and function. Recent systematic reviews, including both observational and interventional studies, support a moderate level of evidence for a role of maternal B12 status in determining offspring cognitive function, and highlight a need for more studies from developing countries (17,30). Studies in Mexico and Singapore have also reported an association between maternal dietary intake of vitamin B12 and offspring cognitive abilities (31,32). Observations in the ALSPAC cohort in the UK suggests a weak association of maternal a genetic determinant of circulating vitamin B12 concentrations (*FUT*2) and offspring IQ at 8 years of age (33). On the other hand, a cohort study in Canada, showed no significant associations between maternal vitamin B12 concentrations and BSID-III outcomes in their offspring at 18 months (32). This may be due to a lack of significant variation in maternal vitamin B12 status, given the low prevalence of vitamin B12 deficiency in their population (34).

Our findings from this pre-conceptional maternal micronutrient supplementation trial fills an important gap in the literature. Our observations are supported by a maternal B12 supplementation study from south India, which supplemented mothers with 50 µg vitamin B12 from the 1^st^ trimester of pregnancy until 6 weeks postpartum. Supplementation improved maternal B12 levels in the third trimester (20) and offspring showed a better neurodevelopmental outcome (language domain) at 30 months of age (21). Vitamin B12 (1.8 µg) and/or folic acid (150 µg) supplementation in 6–30-month-old children for a period of 6 months showed improvement in their neurocognitive performance. The B12 alone group showed improvement in gross motor functioning and the B12 + folic acid group in gross motor as well as problem-solving functioning compared to the placebo; folic acid alone had no effect (35).

The wide prevalence of B12 deficiency is unique to the Indian context due to the socio-cultural practice of vegetarianism. Severe absorption defects (i.e. pernicious anaemia) are rare and vitamin B12 deficiency is largely a low dietary intake problem (15–18). This offers a unique opportunity to control a modifiable risk factor at the public health scale to improve neurodevelopment and human capital in the next generation. Our choice of a near-recommended dietary allowance (RDA) dose of B12 (2 µg/day) was based on our earlier studies showing adequate absorption of oral B-12 (36) in this population and the demonstration in a pilot study of improvement in B-12 and homocysteine status after oral supplementation for 1 year (37). In another study of severely B12 deficient girls, (plasma B12 <100 pmol/l) we demonstrated an improvement in haematological parameters and peripheral and autonomic nerve functions after supplementing 2 µg/day of vitamin B12 for 11 months (38). Thus, we believe that our current study fills an important gap to help public health policy to supplement a physiological dose of vitamin B12 to adolescents and reproductive age women to improve not only their own health but also that of the next generation. Being aware of the difficulties of achieving long term compliance with tablet supplementation in relatively asymptomatic individuals, we have recently reported the efficacy of commonly eaten vitamin fortified food items (nutrient bar and yogurt) to achieve better vitamin B12 status (39). All these approaches are usable in the national programmes to improve micronutrient nutrition of children, adolescents and pregnant mothers.

The improved cognitive outcomes were seen specifically in the B12 alone supplemented group and not in the B12+MMN group. TheB12 alone group achieved higher cord B12, holo-TC and BDNF levels compared to the B12+MMN group, despite a similar daily dose of B12. Though we are unsure about the reason for this difference, the possibilities include differences in compliance and absorption. Evidence for the effects of maternal multiple micronutrient supplementation on offspring outcomes is inconsistent. A systematic review from 9 trials (6 of which used UNIMAP micronutrient composition) did not find favourable effects on child mortality, birth size, or offspring cognition (40).

Vitamins B12 and folate participate in the one-carbon metabolism pathway to 1) stimulate synthesis of precursor nucleotides for DNA synthesis, and 2) generate the universal methyl donor S-Adenosyl methionine (SAM) which is involved in methylation of DNA (an important epigenetic mechanism), proteins and lipids and generating neurotransmitters(41,42). These mechanisms are reputedly involved in fetal growth and differentiation and a deficiency or imbalance of these may result in permanent change in the structure and function of developing tissues which may manifest as disorders in later life (‘fetal programming’) (1). We have demonstrated alterations in adiposity and insulin resistance in children whose mothers had an imbalance of these vitamins (low B12 – high folate) during pregnancy (11). Animal studies have shown differences in the expression of neurotrophic factors such as BDNF in the brains of fetuses whose mothers were exposed to low vitamin B12 status (9). In our study, though we did not find significant differences in cord blood BDNF concentrations between intervention groups, the values tended to be higher in the B12 alone group. Further studies are required to understand the utility of cord BDNF levels as a neurodevelopmental marker in human beings.

Neurodevelopment is a dynamic process that involves neurogenesis, neuronal migration, cortical growth and gyrification, starting in early pregnancy and lasting until infancy (first 1000 days). The pre- and periconceptional period is an important window within this broader window because of ‘epigenetic reprogramming’ of the conceptus which happens within 48-72 hours of conception(43). The majority of pregnancies are unplanned, and women approach the healthcare system after this window. Our intervention was specifically started in adolescence to ensure adequate micronutrient stores in the mother from before conception, in time to support gametogenesis, conception, embryogenesis, organogenesis, and placentation. (42,44). The success of pre-conceptional folic acid supplementation in preventing NTDs is well known (23–25). Thus, we propose that the 1000-day window should be expanded to include the preconception period. This would shift the action from the clinic to the community and will fit well into a multitude of adolescent and reproductive age programs across the world.

Additional strengths of our study include a trial within a cohort in which original observations were made, the RCT design which ensured that potential confounders were similarly distributed between allocation groups. For example, home environmental factors that can influence child neurodevelopment - parental education, maternal IQ and socio-economic status-were similar across intervention groups. High rates of participation in the trial, high rates of follow up, and of sample collection at delivery are also noteworthy. Exclusion of women with severe B12 deficiency (<100pM) from a placebo-controlled trial on ethical grounds reduced the power of the study because they and their offspring could have benefited the most with B12 intervention. The COVID pandemic also interfered with our ability to test more children for neurodevelopment and meant that we missed the children of women who became pregnant later and at older ages. Despite these limitations we were able to see the beneficial effects of the intervention. We expect that the performance on the Bayley’s scale, will reflect in neurodevelopment indices at a later age. This will be tested in subsequent follow ups.

## 5.0 Summary and Conclusion

We found that pre-conceptional maternal supplementation with a near RDA dose (2 µg/day) of vitamin B12 exposed their offspring to higher vitamin B12 status peri-conceptionally and during pregnancy. This was associated with better neurodevelopmental performance in the children, in cognitive and language domains, between 24-42 months of age. Our study highlights an important role for maternal vitamin B12 on offspring neurodevelopment. We urge that the first 1000 days window be extended to include the pre-conceptional period. Our findings have strong implications for public health policy to improve vitamin B12 status of young adolescents and reproductive age women in populations with a sizable vitamin B12 deficiency. We foresee benefits of such a policy to many national nutrition programmes in India.

**Figure 1 Consort depicting the recruitment of study participants**

**Figure 2 Bar graph showing comparison between placebo and treatment groups on BSID-III domains**

## Data Availability

Data is available from the corresponding author on a suitable request. Data sharing is subject to KEMHRC Ethics Committee approval and Government of India Health Ministry advisory committee permission.

## Authors contributor statement

CSY and RVB designed the neurocognitive follow-up study. ND and RVB analysed the data and wrote the first draft. BP and MD performed and reported the neurocognitive assessments. DB performed the biochemical measurements. AB contributed to the statistical analysis. SS and RS conducted the follow-up of the participants. KK, RL, and PY contributed to conducting the PRIYA trial. CSY and CF designed the original PRIYA trial. CSY and CF edited the final manuscript. All authors contributed to the article and approved the submitted version.

## Acknowledgments

We are grateful to the participants of the PMNS for their cooperation in this trial. We acknowledge the contribution of the field workers - Mr Ankush Bhalerao, Mr Suresh Chougule, Mr Vishnu Solat and Ms Rajani Bendge, Ms Malti Raut, Ms Mangal Gaikwad, Ms Sujata Jagtap, Ms Sarla Bagate, Ms Jayshri Kalokhe in conducting the trial. We thank Ms Anagha Deshmukh, Ms Sarah Khan, Ms Deepa Raut, Ms Sayali Wadke, Ms Rajashree Kamat for their assistance in data collection.

