## supplementary tables for "Pre-conceptional maternal vitamin B12 supplementation improves offspring neurodevelopment at 2 years of age: PRIYA trial"

**Supplementary Table 1.** Maternal characteristics and child characteristics of assessed group compared to non-assessed group

| **Variables** | **n** | **Assessed group** | **n** | **Not assessed group** | **P value** |
| --- | --- | --- | --- | --- | --- |
| **Parental sociodemographic characteristics** | | | | | |
| Maternal age at 28 weeks gestation (years) | 82 | 19.6 (18.5, 20.3) | 62 | 22.5 (21.6, 23.0) | <0.001*** |
| Maternal education (years) | 84 | 12.0 (10.0, 13.0) | 62 | 15.0 (12.0, 15.0) | <0.001*** |
| Maternal height (cms) | 82 | 157.0 (5.1) | 61 | 156.0 (4.8) | 0.067 |
| Maternal weight at 28 weeks gestation (kgs) | 82 | 52.1 (48.4, 57.7) | 61 | 55.1 (50.1, 60.5) | 0.915 |
| Maternal IQ | 41 | 75.0 (71.0, 81.0) | 19 | 81.0 (79.0, 92.0) | 0.002** |
| Standard of Living Index | 84 | 36.5 (31.2, 40.0) | 62 | 36.5 (31.0, 40.2) | 0.763 |
| Paternal Education (years) | 84 | 12.0 (10.0, 15) | 62 | 15 (12, 15) | 0.006** |
| **Maternal Micronutrients** | | | | | |
| Maternal B12 at screening (pM) | 85 | 148 (125, 202) | 64 | 147 (127, 187) | 0.677 |
| Maternal Folate (nM) at screening | 85 | 20.9 (13.8, 27.1) | 64 | 18.5 (15.0, 25.3) | 0.965 |
| Maternal Homocysteine (µmol/L) at screening | 85 | 21.1 (16.4, 32.4) | 64 | 22.0 (17.9, 33.8) | 0.416 |
| Maternal B12 at 18 years (pM) | 76 | 225 (162, 326) | 61 | 226 (152, 287) | 0.861 |
| Maternal Folate (nM) at 18 years | 78 | 21.2 (15.8, 29.7) | 61 | 23.2 (18.1, 31.0) | 0.130 |
| Maternal Homocysteine (µmol/L) at 18 years | 79 | 11.8 (9.30, 16.8) | 61 | 11.5 (9.60, 18.7) | 0.583 |
| **Maternal Micronutrient levels at 28 weeks gestation** | | | | | |
| Hemoglobin (gm/dl) | 82 | 10.4 (9.40, 11.0) | 61 | 10.6 (9.80, 11.6) | 0.078 |
| B12 (pM) | 82 | 166 (126, 235) | 61 | 210 (156, 305) | 0.005** |
| Holo-TC (pM) | 82 | 19.6 (12.5, 31.0) | 61 | 34.7 (20.0, 89.3) | <0.001** |
| Folate (nM) | 82 | 31.4 (16.2, 61.1) | 61 | 45.6 (24.0, 60.2) | 0.122 |
| B2 (pM) | 81 | 243.0 (221.0, 275.5) | 61 | 198.0 (175.5, 238.0) | <0.001*** |
| B6-pyridoxal-5-phospate (pM) | 81 | 3.80 (2.90, 5.50) | 61 | 4.10 (2.90, 5.60) | 0.572 |
| B6-pyridoxal (pM) | 81 | 1.20 (0.94, 1.60) | 61 | 1.60 (1.20, 2.30) | <0.001*** |
| Homocysteine (µmol/L) | 82 | 6.60 (4.27, 8.20) | 61 | 6.60 (5.20, 8.95) | 0.281 |
| **Child Characteristics** | | | | | |
| Gender | 85 | Boys= 48 (56.5%) | 63 | Boys= 31 (49.2%) |  |
| **Birth Anthropometry** | | | | | |
| Birth weight (gms) | 85 | 2700 (2377, 3000) | 64 | 2780 (2545, 3015) | 0.338 |
| Birth length (cms) | 84 | 48.2 (47.0, 49.2) | 60 | 48.2 (47.1, 49.1) | 0.943 |
| Head circumference (cms) | 84 | 33.1 (32.4, 33.9) | 60 | 33.5 (32.8, 34.3) | 0.020 |
| Gestation age (months) | 85 | 39.2 (38.2, 40.1) | 64 | 39.5 (38.7, 40.1) | 0.562 |
| **Cord Micronutrient** | | | | | |
| B12 (pM) | 85 | 243 (165, 373) | 58 | 369 (223, 808) | 0.001** |
| Holo-TC (pM) | 85 | 61.3 (27.5, 119) | 40 | 121 (47.2, 128) | 0.004** |
| Folate (nM) | 85 | 9.30 (6.55, 15.5) | 57 | 10.3 (7.45, 13.8) | 0.730 |
| B2 (pM) | 81 | 321 (263, 379) | 58 | 274 (242, 305) | <0.001*** |
| B6-pyridoxal-5-phospate (pM) | 85 | 26.0 (15.8, 37.6) | 58 | 27.4 (17.2, 38.7) | 0.557 |
| B6-pyridoxal (pM) | 85 | 4.90 (3.75, 7.15) | 58 | 4.50 (3.47, 6.10) | 0.138 |
| Homocysteine (µmol/L) | 85 | 7.40 (5.00, 10.7) | 58 | 6.20 (4.25, 8.20) | 0.020* |

*p<0.05, **p<0.01, ***p<0.001 p-values calculated by students t-test

Values represented as Mean (SD) Median (25th, 75th) or n (%)

Holo-TC, holotranscobalamin; BDNF, Brain Derived Neurotrophic Factor

**Supplementary Table 2.** Comparison of BSID performance across treatment groups

| **BSID-III domains** | **Category** | **Placebo** | **B12+MMN** | **B12** | **Chi-square**  **p value** |
| --- | --- | --- | --- | --- | --- |
| Cognitive | Above average | 0% | 3.8% | 9.5% | 0.480 |
|  | Average | 96.3% | 92.3% | 90.5% |  |
|  | Below average | 3.7% | 3.8% | 0% |  |
| Motor | Above average | 7.4% | 11.5% | 9.5% | 0.904 |
|  | Average | 81.5% | 84.6% | 90.5% |  |
|  | Below average | 3.7% | 3.8% | 0% |  |
| Language | Above average | % | 3.8% | 14.3% | 0.241 |
|  | Average | 92.6% | 88.5% | 76.2% |  |
|  | Below average | 3.7% | 7.7% | 4.8% |  |

Values represented as Mean (SD) or Median (Q1, Q3)

p-values calculated by chi-square

**Supplementary** **Table 3.** Comparison of BSID performance between the sexes

| BSID-III domains | Male | Female | p value |
| --- | --- | --- | --- |
| Cognitive | 90.0 (85.0, 95.0) | 95.0 (90.0, 100) | 0.099 |
| Motor | 94.0 (89.5, 100) | 97 (91.0, 107) | 0.329 |
| Language | 93.2 (10.4) | 96.4 (7.71) | 0.160 |

Values represented as Mean (SD) or Median (Q1, Q3)

p-values calculated by t-test

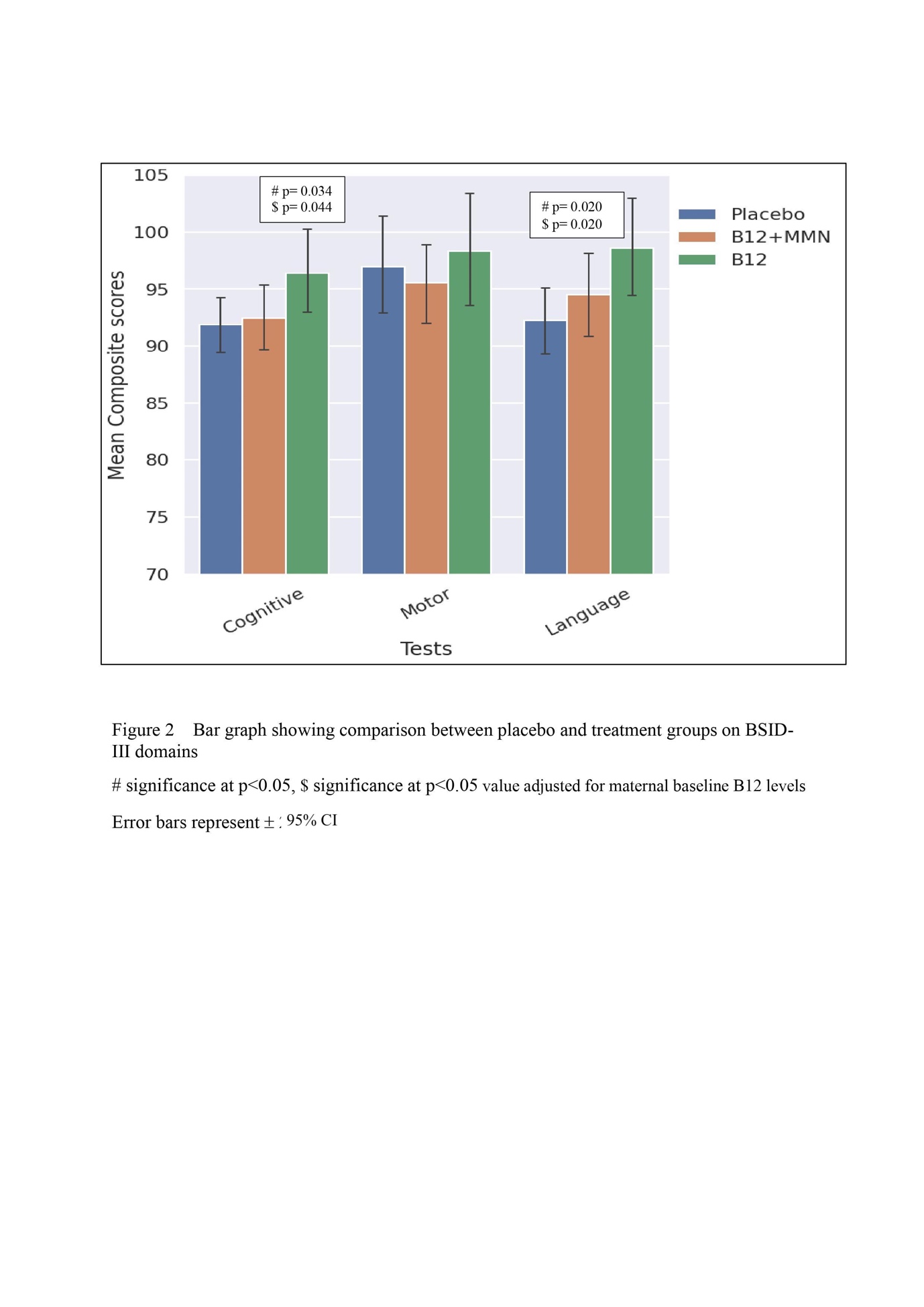
